# Isolation and Evaluation of a Lytic Staphylophage With Activity Against Methicillin Resistant *Staphylococcus Aureus* from Postsurgical Wound Infections in Surgical Wards at Mzuzu Central Hospital, Malawi

**DOI:** 10.64898/2026.09.21.26354812

**Authors:** Evelyn Mwangomba, Rashid Kaseka, Patricia Bazwell Banda, Gama Bandawe

**Affiliations:** MUST Bacteriophage Research Unit, Center for Clinical and Biological Sciences Research, Academy of Medical Sciences, Malawi University of Science and Technology, Ndata Campus, Malowa-Goliati S150 Road, T/A Chimaliro, 310105, Thyolo, Malawi; Mzuzu Central Hospital, Private Bag 209, Luwinga, Mzuzu 2, Malawi; Frankfurt University Hospital, Theodor-Stern-Kai 7, 60596 Frankfurt am Main, Germany

## Abstract

**Background:** Methicillin-resistant *Staphylococcus aureus* (MRSA) is one the most common cause of post-surgical infections in Malawi and poses a significant public health challenge due to resistance to commonly used antibiotics. We isolated and evaluated the stability and lytic activity of a MRSA-specific bacteriophage (Staphylophage) from Mzuzu Central hospital (MZCH) sewage against MRSA strains isolated from surgical wards.

**Methods:** A prospective in vitro experimental study was conducted using 56 pus swabs collected from post-surgical patients at Mzuzu Central Hospital (MZCH), Malawi. Standard microbiological methods and VITEK analysis were used for bacterial isolation, identification and antimicrobial susceptibility testing of *Staphylococcus aureus* and MRSA according to the 2024 EUCAST guidelines. A lytic Staphylophage was isolated and purified from sewage wastewater using filtration, enrichment, and double-layer agar techniques. The purified Staphylophage was evaluated for its lytic activity through host range testing and stability under ultraviolet (UV) exposure (up to 60 minutes), different pH levels (2–12), and temperatures (4°C, −20°C, and −80°C).

**Results:** Out of 56 bacterial isolates, 18 (32.1%) were *S. aureus*, of which 11 (61.1%) were MRSA. The isolated Staphylophage showed lytic activity against 87% of *Staphylococcus* species tested. The phage remained stable at temperatures between −80°C and 4°C and across a pH range of 2– 12. It also retained viability after UV exposure for up to 15 minutes, although a reduction in phage titre was observed after 30 minutes.

**Conclusions:** A lytic Staphylophage isolated from hospital sewage at MZCH demonstrated activity against *Staphylococcus* species, including MRSA strains from post-surgical wound infections. These findings illustrate the feasibility of generating candidate phages for alternative therapeutic approaches in a central hospital setting in Malawi.

## INTRODUCTION

The rapid rise of antimicrobial resistance (AMR) has significantly reduced the effectiveness of many conventional antibiotics. This challenge is particularly associated with the ESKAPE group of pathogens—*Enterococcus faecium, Staphylococcus aureus, Klebsiella pneumoniae, Acinetobacter baumannii, Pseudomonas aeruginosa,* and *Enterobacter* species—which are responsible for a large proportion of multidrug-resistant infections worldwide (1,2). Among these organisms, Methicillin-Resistant *Staphylococcus aureus* (MRSA) has emerged as a major public health concern due to its resistance to β-lactam antibiotics and its association with prolonged hospitalization, increased morbidity, mortality, and healthcare costs (3). Globally, MRSA is a leading cause of hospital-acquired infections, particularly in surgical site infections, bloodstream infections, and skin and soft tissue infections (4–7).

The burden of MRSA is particularly significant in Africa, where healthcare systems face challenges related to limited diagnostic capacity, poor infection prevention practices, limited microbiological surveillance, inadequate antimicrobial stewardship programs, and restricted access to effective antimicrobial agents. Prevalence of MRSA has been reported at approximately 36.7% across Sub-Saharan Africa, (8) and 39.9% within Eastern Africa, with considerable variation exists between countries (9).

In Malawi, *Staphylococcus aureus* remains one of the most frequently isolated bacterial pathogens in clinical settings (10). with a significant portion of these being identified at MRSA. Studies conducted at Malawi’s major referral hospitals have reported high or increasing MRSA prevalence. At Kamuzu Central Hospital 31.3% of all *S. aureus* isolates were identified as MRSA (1), while at Queen Elizabeth Central Hospital, an analysis of blood culture isolates reposted a prevalence of 9.6% between 1998 and 2016 followed by an elevated prevalence of 10.5% between and (11).

In surgical wards where S. aureus is the main source of infection in post-operative wounds, MRSA infections remain a dire and persistent challenge. Analysis of WHONET surveillance data from 2021–2023 from referral hospitals showed that Mzuzu Central Hospital (MZCH) had the highest incidence of MRSA among postsurgical patients at 22% compared to Kamuzu Central hospital (20%) and QECH (15%). The rising incidence of MRSA, coupled with dwindling treatment options, calls for immediate exploration of alternative antimicrobial strategies such as bacteriophage therapy for which there is growing interest globally (12).

Environmental reservoirs such as hospital wastewater have been identified as rich sources of Staphylophage capable of targeting clinically relevant MRSA bacterial pathogens. Unlike broad-spectrum antibiotics, phages are highly specific to their bacterial hosts, which reduces off-target effects and preserves beneficial microbiota. While phage therapy has seen a resurgence in parts of Europe and Asia, its application remains very limited and largely experimental in Africa for numerous reasons including lack of capacity and resources. Some African investigators have successfully isolated MRSA phages from hospital wastewater, with activity of 80% lysis rates against clinical MRSA isolates (13). However, to date, no published study in Malawi on MRSA phages are available and bacteriophage research in the country is still in its infancy as the country faces significant gaps in local phage research and application. To advance the development of locally relevant alternative antimicrobial interventions we sought to isolate and evaluate highly active Staphylophage from local wastewater sources and assess their lytic activity against clinical MRSA isolates obtained from surgical wound infections at Mzuzu Central Hospital

## METHODS

### Study Location

The study was conducted at Mzuzu Central Hospital, a tertiary referral hospital in the Northern Region of Malawi catering for a population of two million five hundred thousand people with a bed capacity of 384.

### Methodological approach

A summarized methodological view of the study is depicted in figure 1 below.

**Figure 1.**
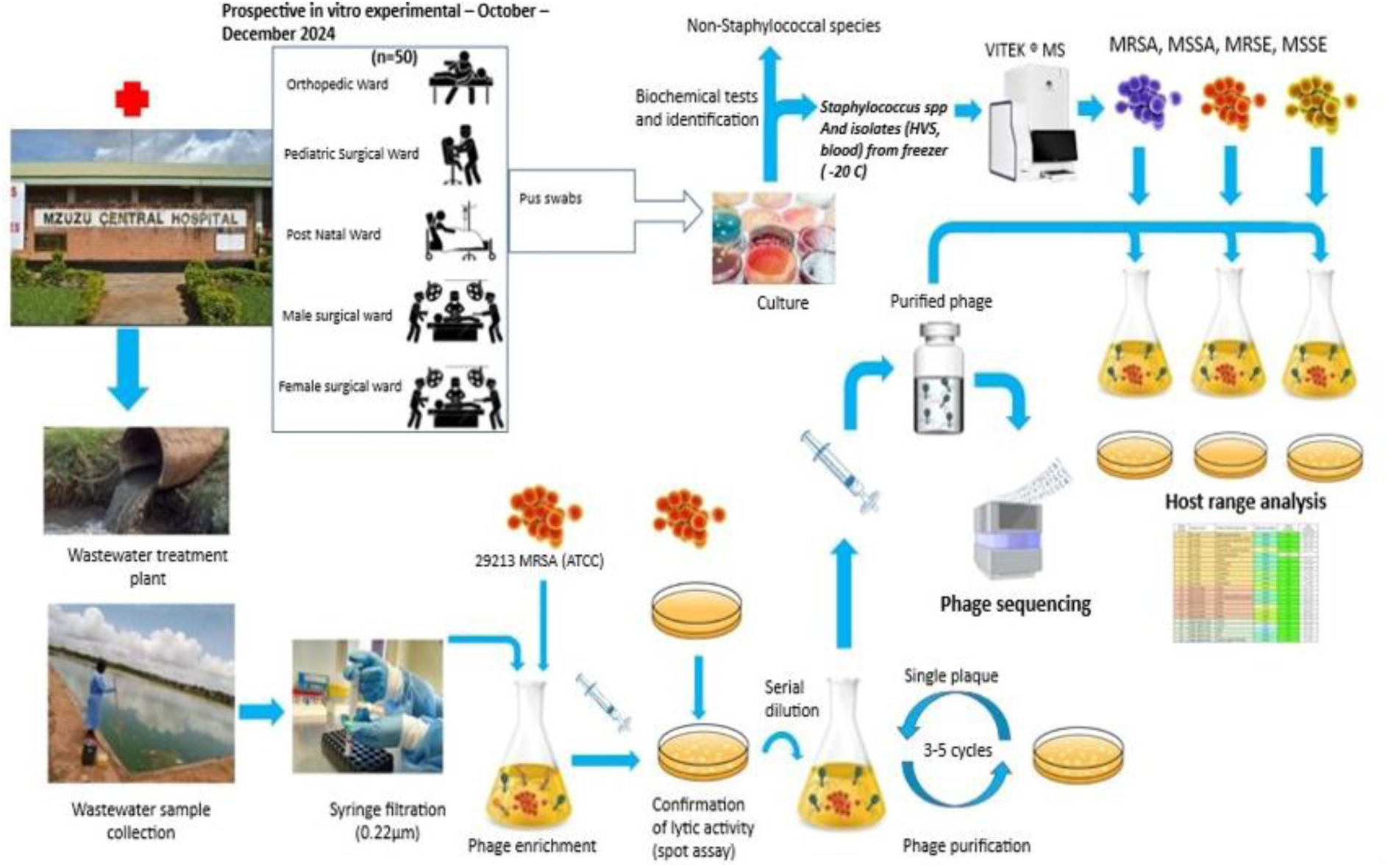
Schematic overview of the study design for isolation, characterization, and evaluation of Staphylophage. Clinical pus swab samples (n = 50) were collected from patients admitted to the orthopedic, pediatric surgical, postnatal, male surgical, and female surgical wards at Mzuzu Central Hospital between October and December 2024. Staphylococcal isolates obtained from clinical samples and archived isolates were cultured, identified using biochemical tests and VITEK® MS, and categorized as methicillin-resistant or methicillin-susceptible Staphylococcus aureus (MRSA, MSSA) and Staphylococcus epidermidis (MRSE, MSSE). Wastewater samples collected from a wastewater treatment plant were filtered through a 0.22 μm membrane and enriched using the reference MRSA strain ATCC 29213 to isolate lytic bacteriophages. Phage activity was confirmed by spot assays, followed by serial dilution and plaque purification through 3–5 rounds of single-plaque isolation. Purified phages were subsequently sequenced and evaluated for host range against the panel of clinical staphylococcal isolates to determine their lytic spectrum.

### Culture Media Quality Control

All culture media used in this study, including Blood Agar (BA-BD), Mueller–Hinton Agar (MHA-BD), agarose, and Tryptone Soy Broth (TSB) (BD Diagnostics, USA), were subjected to quality control procedures prior to use. Media sterility and performance testing were conducted following standard microbiological quality control guidelines.

To assess sterility, randomly selected prepared plates, broth tubes and incubated at 35–37 °C for 48 hours without inoculation. The absence of microbial growth after incubation confirmed that the media preparation procedures were sterile.

Media viability and performance were evaluated using a reference control strain, *Staphylococcus aureus* ATCC 29213. The organism was inoculated onto the prepared media and incubated under appropriate conditions. Successful growth of the control organism confirmed that the culture media supported the growth of fastidious and common microorganisms and allowed detection of characteristic hemolytic activity on blood agar.

### Sample Collection and Processing for MRSA isolation

Fifty (50) admitted post-surgical patients who developed wound infections 48 hours after surgery or up to 90 days following surgery involving implants were included in this study. Pus swab samples were collected from patients presenting with post-surgical wound infections in various surgical wards at Mzuzu Central Hospital, including the orthopedic ward, pediatric surgical ward, male surgical ward, and female surgical ward. Sample collection was conducted between October and December 2025.

Sterile cotton swabs were used to collect pus specimens aseptically from infected surgical wounds. The swabs were immediately placed in Amies Stuart transport medium and transported promptly to the microbiology laboratory to maintain sample integrity. Samples that could not be processed immediately were stored at 4 °C to preserve bacterial viability until laboratory analysis.

In the laboratory, all procedures were conducted under Biosafety Level II (BSL-II) conditions. Each sample was suspended in 5 mL of sterile normal saline and mixed thoroughly to ensure uniform distribution of microorganisms. A sterile loop or swab was then used to streak a portion of the suspension onto blood Agar (BA) plates using the three-quadrant streaking technique to obtain isolated colonies.

The inoculated plates were incubated at 37 °**C** for 24–48 hours under aerobic conditions. After incubation, colonies were examined for morphological characteristics typical of *Staphylococcus aureus*. Presumptive *S. aureus* colonies appeared large, round, smooth, convex, opaque, and golden-yellow in color, with or without β-hemolysis on blood agar.

### Identification and Confirmation of *Staphylococcus aureus*

Colonies suspected to be *Staphylococcus aureus* based on morphology on Blood Agar were further subjected to a series of standard microbiological identification tests. Initially, **Gram staining** was performed to determine cell morphology and Gram reaction. Smears prepared from isolated colonies were heat-fixed, stained, and examined microscopically. Gram-positive cocci appearing in grape-like clusters were considered presumptive *Staphylococcus* species.

Presumptive isolates were further tested using the **catalase test** to differentiate staphylococci from streptococci. A small portion of the colony was placed on a clean glass slide and a drop of 3% hydrogen peroxide was added. The immediate production of bubbles indicated a positive catalase reaction.

Catalase-positive isolates were subsequently subjected to the **coagulase test** for confirmation of *Staphylococcus aureus*. The slide coagulase test was first performed by emulsifying a colony in a drop of sterile saline on a glass slide, followed by the addition of plasma. The formation of visible clumping within a few seconds indicated a positive reaction. Isolates that produced positive results were considered confirmed *Staphylococcus aureus*. When necessary, the tube coagulase test was used to confirm borderline or negative slide test results by incubating bacterial suspensions with plasma at 37 °C and observing clot formation after 4–24 hours.

### Detection and Confirmation of Methicillin-Resistant *Staphylococcus aureus* (MRSA)

Confirmed *Staphylococcus aureus* isolates were screened for methicillin resistance using the **cefoxitin disk diffusion method** in accordance with recommendations provided by the **European Committee on Antimicrobial Susceptibility Testing (EUCAST)**.

A bacterial suspension equivalent to 0.5 McFarland turbidity standard was prepared from freshly grown colonies. The suspension was evenly inoculated onto Mueller–Hinton agar plates using a sterile cotton swab to obtain a uniform bacterial lawn. A thirty microgram (30 µg) cefoxitin disk was placed onto the inoculated agar surface, and the plates were incubated aerobically at 35–37 °C for 18–24 hours (14).

Following incubation, the diameter of the inhibition zone surrounding the cefoxitin disk was measured in millimetres (mm). According to EUCAST interpretive criteria, isolates showing reduced cefoxitin disc zone diameter (≤ 22mm) were considered potential MRSA, while those exhibiting larger zones of inhibition (≥ 22mm) were classified as methicillin-susceptible *Staphylococcus aureus* (MSSA).

For further confirmation, isolates identified as MRSA plus additional randomly selected 5 % of the stored (-20 ° C) isolates (composed of new isolates from pus swabs, blood and High vaginal swabs) were sent for MRSA confirmation at Public Health Institute of Malawi (PHIM) laboratory. At PHIM the isolates were further analysed using the automated **VITEK 2 System** (bioMérieux), for microbial identification and antimicrobial susceptibility testing (15). Pure colonies were suspended in sterile saline and adjusted to the required turbidity level (0.5 Mcfarland) following the manufacturer’s instructions. The suspension was inoculated into identification and antimicrobial susceptibility testing cards specific for Gram-positive organisms (*Staphylococcus species*). The cards were then loaded into the VITEK 2 instrument for automated analysis. The VITEK 2 system generated results based on biochemical reactions and antimicrobial susceptibility profiles, thereby confirming the identification of *Staphylococcus aureus* with Minimum Inhibitory Concentration (MIC) values of greater than four milligram per litre (> 4 mg/l) were determined as methicillin resistance(16)

Quality control for antimicrobial susceptibility testing was performed using the reference strain Staphylococcus aureus ATCC 25923 to ensure accuracy and reliability of the results.

### Isolation of Bacteriophages

Bacteriophages were isolated from raw sewage samples collected from the sewage treatment points at Mzuzu Central Hospital (MZCH), following a standard operating protocol developed for phage isolation. The protocol was developed from previous similar protocols to suit the limited resources and environment (17).

Upon sample receipt, 50 mL falcon tubes containing the sewage water were labeled and balanced in a refrigerated centrifuge at 4°C. They were centrifuged at 4000 rpm for 30 minutes to remove large debris. The resulting supernatant was carefully aspirated using a sterile syringe and filtered through a 0.22 µm PES membrane syringe filter into a second sterile falcon tube. Filtrates were stored at 4°C until used for phage enrichment and testing.

For enrichment, 200 µL of the filtrate was mixed with 200 µL of a 6-hour culture of the target host bacteria—Methicillin-Resistant *Staphylococcus aureus* (MRSA), grown in 4 mL Tryptone Soy Broth (TSB). This mixture was added to 3.5 mL of soft agar (TSB + 6% agarose) and poured onto solidified Tryptic Soy Agar (TSA) bottom agar plates. The overlay was gently swirled and allowed to solidify in biosafety level 2 cabinet (BSLII) approximately 20 minutes to reduce contamination risk. Plates were incubated aerobically at 37°C for 24 hours. Following incubation, plates were examined for clear lysis zones (plaques), which were taken as confirmation of lytic phage activity against the bacterial host. Positive lysates were noted for further host range and stability testing. The inclusion of multiple target bacterial species (MRSA, *K. pneumoniae*, and *E. coli*) aimed to test specificity and broaden the detection and potential therapeutic use of sewage-derived phages in combating resistant pathogens within hospital environments.

### Purification of Bacteriophages

Following initial isolation and confirmation of lytic activity, phage lysates were subjected to a purification process to obtain clonal, high-titer phage stocks for downstream analysis and testing (18).

### Plaque Picking and Enrichment

Distinct plaques (clear zones) with similar morphology were aseptically picked from TSA plates using a sterile disposable loop and suspended in 1 mL of SM buffer within a sterile 50 mL conical tube. Suspensions were stored at 4°C to maintain phage viability. An aliquot of 100 µL of the plaque suspension was inoculated into 4 mL of Tryptone Soy Broth (TSB) containing a 6-hour-old culture of the corresponding host MRSA bacterium, and incubated at 37°C for 6 hours to enrich active phages(18).

### Centrifugation and Filtration

Post-incubation, the mixture was centrifuged at 4000 rpm for 30 minutes using a refrigerated centrifuge at 4°C. The resulting supernatant was carefully separated from cell debris and filtered using a 10 mL syringe fitted with a 0.22 µm PES membrane filter into sterile tubes to obtain purified lysate.

### Serial Dilution and Re-plating

The filtered lysate was serially diluted tenfold from 10⁻¹ to 10⁻⁷ in SM buffer. Initially each of 7 tubes were aliquoted with 900 µL of SM buffer mixed with 100 µL of lysate plus 100 µL of a fresh 6-hour-old host culture while subsequent tubes were aliquoted by taking 100 µL from the first tube and incubated at 37°C for 3 hours to facilitate phage adsorption. Double-layer agar plates were prepared with a bottom layer of TSA (TSB + 6% agarose) and a top layer of TSB with 0.7% agarose. Top agar was maintained in a water bath at 50°C until use. For each dilution, 10 µL of the phage-host mixture was spotted onto the surface of the top agar, allowed to solidify for 30 minutes, and incubated overnight at 37°C. Plates were examined for individual plaques in the spots. Plaques from the highest dilution with distinct morphology were re-picked and suspended in 500 µL of SM buffer, followed by centrifugation at 4,000 rpm for 30 minutes. The supernatant was filtered again through a 0.22 µm filter. This purification cycle was repeated 3–5 times to ensure clonal phage isolation.

### Phage Titer Determination and Final Bulk-Up and Storage

The final phage titer was calculated using the following formula: PFU/mL = Number of plaques per plate ÷ Dilution factor × Volume plated (mL). Calculating phage titer requires a purified phage plate. This standardized purification step ensured the production of stable, concentrated phage stocks with known titers for downstream stability testing and host range assays. A high-titer stock was produced by scooping similar plaque morphology to yield purified phage. A 10 µL of lysate was mixed with 100 µL of fresh host culture, aliquoted into 3.5 mL of top agar and poured on TSA bottom agar. After solidification and 24-hour incubation at 37°C, 5 mL of SM buffer was added to the plate and incubated at 4°C in the refrigerator overnight. The buffer containing phages was recovered using a syringe and filtered again through a 0.22 µm filter into sterile 5 mL tubes.

### Data Management and Statistical Analysis

All data generated during the study were systematically recorded, cleaned, and validated prior to analysis. Clinical and microbiological data, including patient demographics, isolate identification, and antimicrobial susceptibility results, were initially captured using WHONET, a standardized software developed by the World Health Organization for the management and analysis of antimicrobial resistance surveillance data (19).

Subsequently, the cleaned datasets were exported to Microsoft Excel for data organization and preprocessing. Statistical analyses were conducted using Python (version 3.x) and R statistical software (version 4.3.1). Data were analyzed descriptively. Proportions were used to summarize bacterial isolates and MRSA prevalence. Phage activity and stability results were reported based on observed lytic activity and changes in phage titers under different conditions.

In this study, phage activity (PFU/mL) was measured across a variety of experimental conditions including storage temperatures, pH levels, and UV exposure durations. Due to logistical and resource constraints during laboratory experimentation, most conditions were assessed using single measurements without technical or biological replicates. Consequently, it was not possible to compute standard deviations or include error bars in the graphical representations. While this limits the ability to quantify variability. Statistical confidence across replicates, the results still provide useful exploratory insights into the trends and comparative effects of these stress conditions on phage viability.

## RESULTS

### Prevalence of Staphylococcus aureus and MRSA in Surgical Wards

Among the 56 total clinical isolates obtained from post-surgical wound samples from 50 participants, *Staphylococcus aureus* was the most frequently isolated organism, accounting for 18 isolates (32.1%) as summarized in table 1 below. Of these, 11 isolates (61.1%) were confirmed as methicillin-resistant (MRSA) using the cefoxitin disk diffusion method, in line with EUCAST (2024) standards.

**Table 1:**
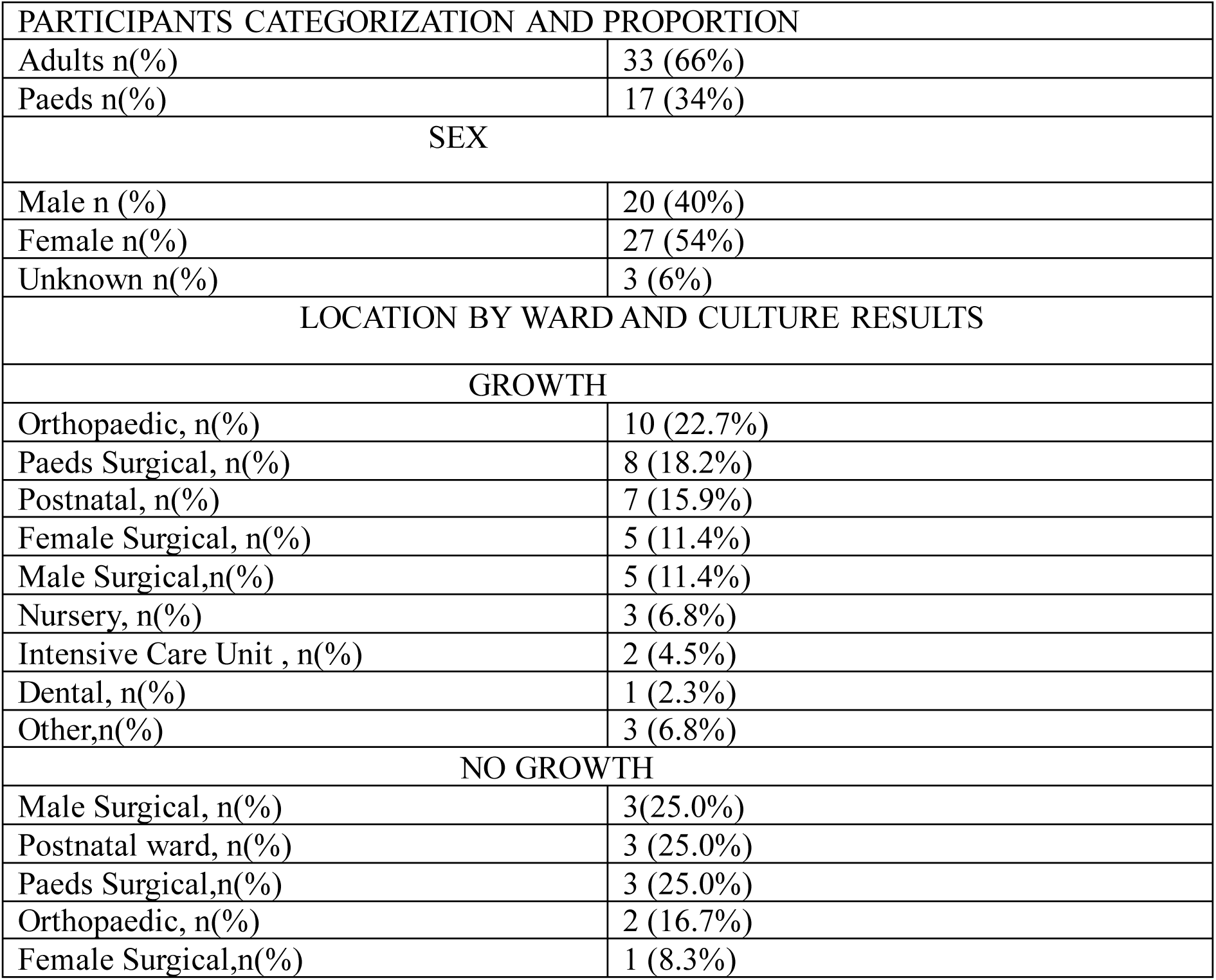
Demographic characteristics of participants. Data are presented as frequency and percentage, *n (%)*. A total of 50 participants (56 isolates) were included, comprising 36 adults (64.3%) and 20 paediatric patients (35.7%). Sex distribution was 21 males (42%), 26 females (52%), and 3 participants with unknown sex (6%). Culture results were categorized as growth and no growth. Among specimens with culture growth, the highest proportions were from the Orthopaedic ward (22.7%), Paediatric Surgical ward (18.2%), and Postnatal ward (15.9%). Among specimens with no growth, the highest proportions were from the Male Surgical, Postnatal, and Paediatric Surgical wards, each contributing 25.0% of no-growth cultures. Percentages were calculated within each respective category (participant characteristics and culture result groups).

**Figure 2.**
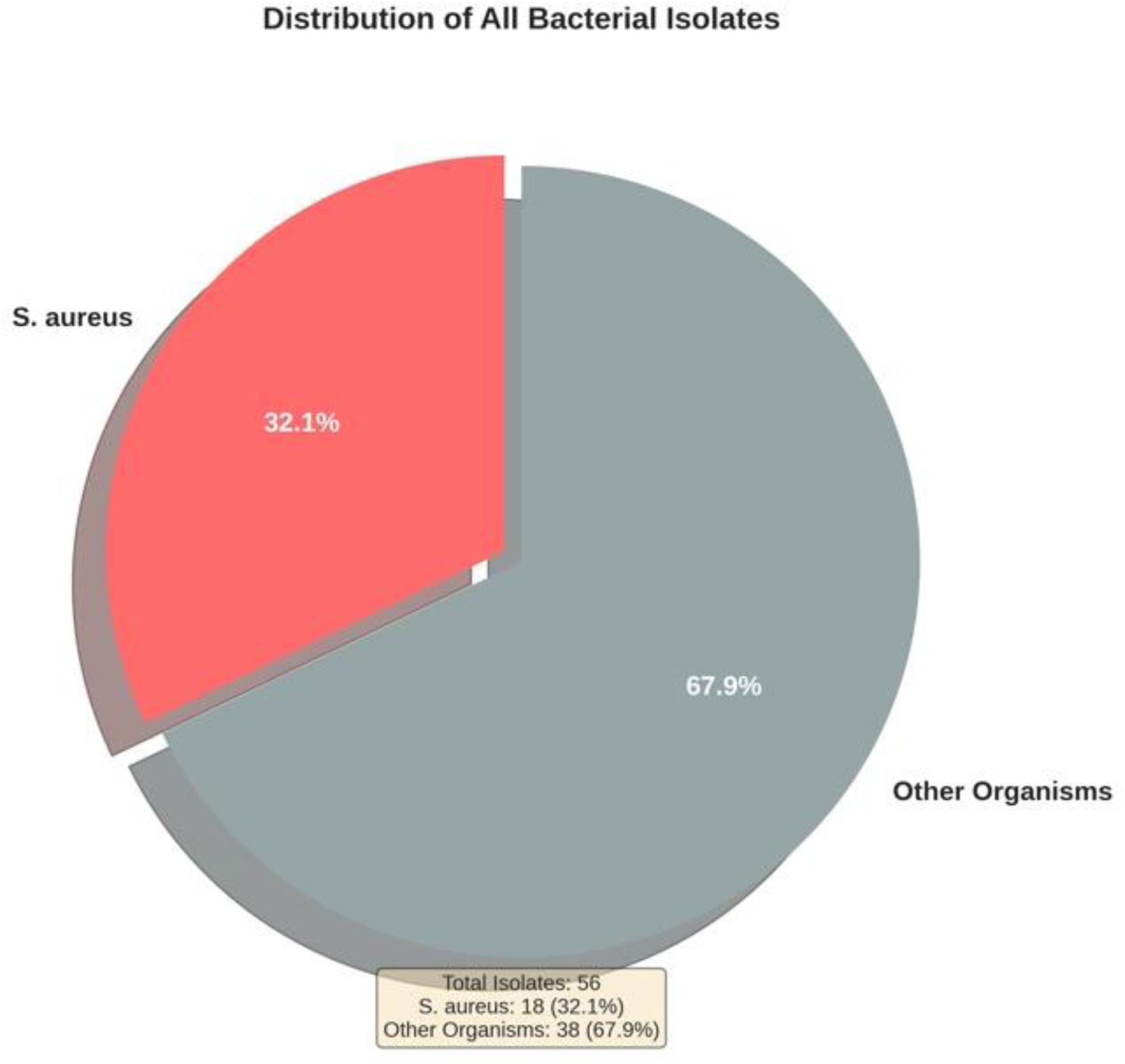
Distribution of bacterial isolates recovered from clinical specimens. A total of 56 bacterial isolates were obtained from clinical samples. Staphylococcus aureus accounted for 18 isolates (32.1%), while other bacterial species comprised 38 isolates (67.9%). The pie chart illustrates the relative proportion of S. aureus compared with all other recovered organisms.

**Figure 3.**
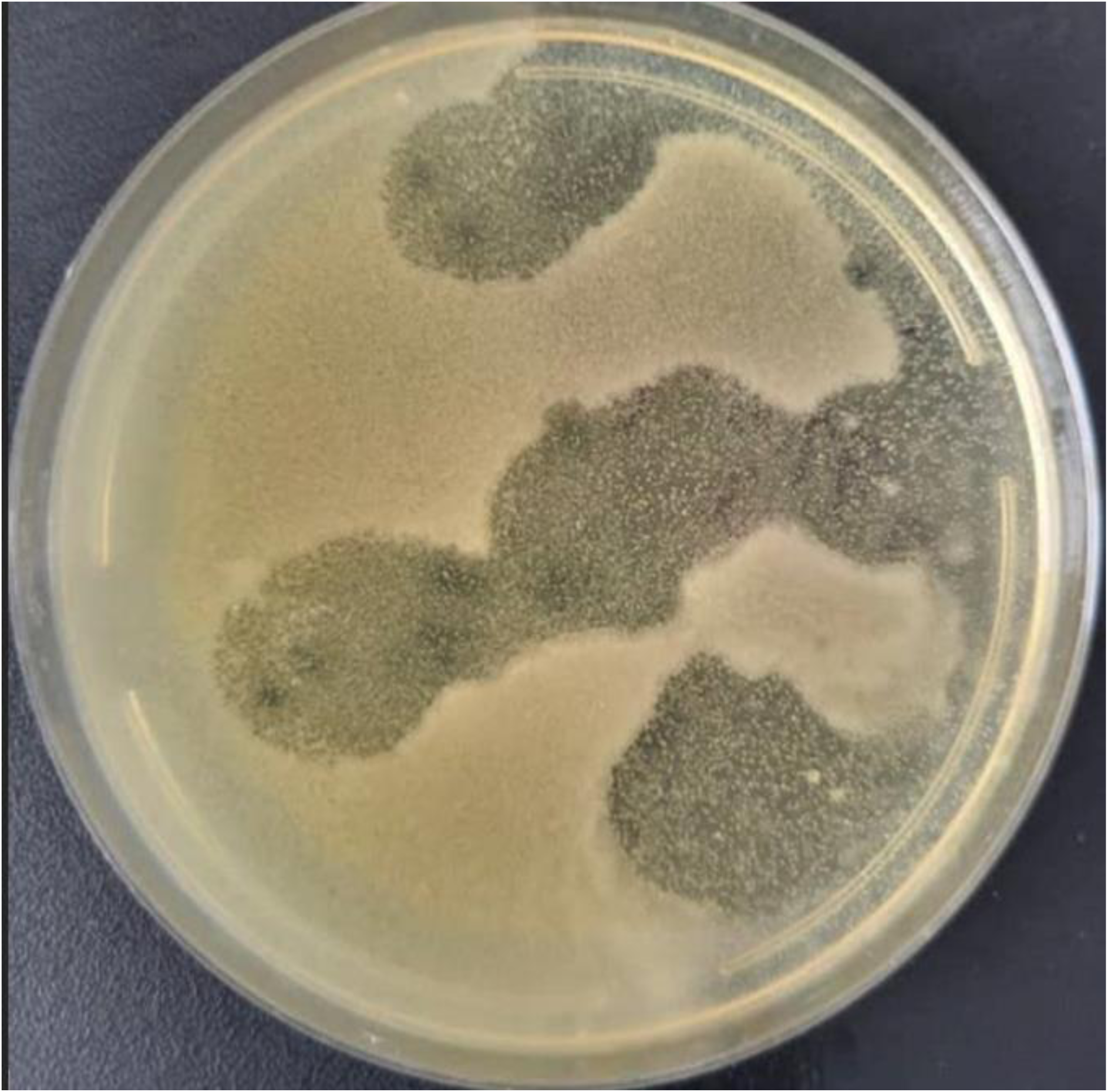
Representative Spot assay performed using filtered and enriched wastewater. showing lytic activity of Staphylophage against Staphylococcus aureus ATCC 29213. Muiltiple 10 µL phage lysate spots were applied to confluent to bacterial lawn Trypton Soy Agar (TSA-BD) and incubated at 37°C for 24 h. Clear lytic zones developed at the sites of phage application indicating efficient adsorption, replication and host cell lysis. Upon visual inspection of plaque morphology on the positive plate, the plaques appeared dissimilar in size and shape, suggesting that more than one phage type, or unrelated phages with different lytic patterns, were present.

The study isolated pure Staphylophage using *Staphylococcus aureus* American Type Culture Collection (ATCC 29213) as positive control to purify and aid visualization as indicated in **figure 4** below.

**Figure 4.**
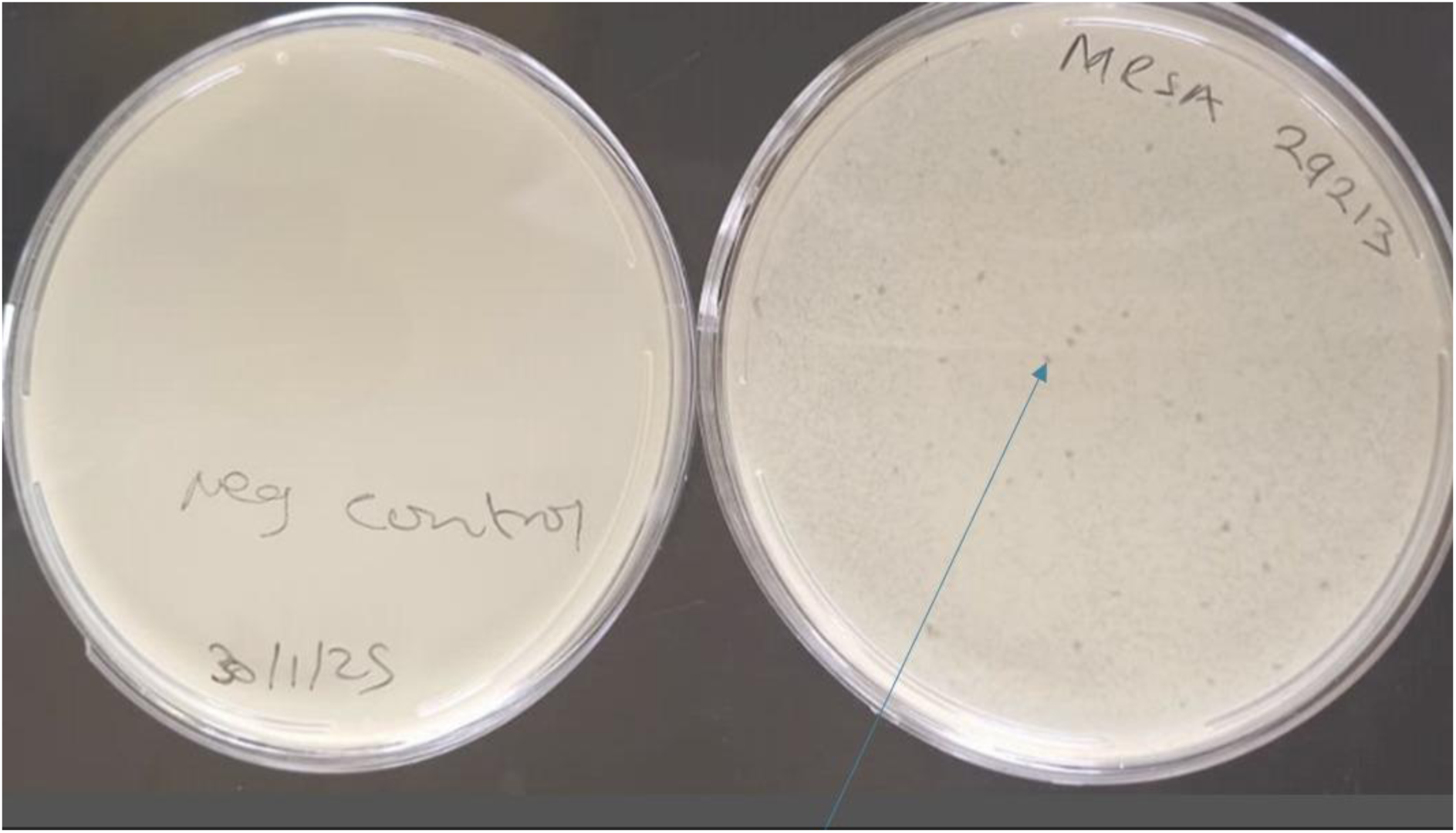
Fourth-round plaque purification of staphylophage using Staphylococcus aureus ATCC 29213 as the host strain. The left plate represents the negative control and shows no evidence of lysis or contamination. The right plate shows a bacterial lawn of S. aureus ATCC 29213 on tryptic soy agar (TSA) following the fourth round of plaque purification. Well-isolated plaques are visible throughout the lawn, indicating successful infection and lysis by individual phage particles. A single plaque was selected from this plate for subsequent propagation to ensure the establishment of a genetically homogeneous Staphylophage population. Plates were incubated at 37°C for 24 h prior to imaging.

### Host range determination of the isolated phage

To determine the clinical spectrum of activity of the isolated phage, phage lysis assays were conducted on a panel of *Staphylococcus species* isolates namely: Methicillin Resistant *Staphylococcus aureus* (MRSA), Methicillin Susceptible *Staphylococcus aureus* (MSSA), Methicillin Resistant *Staphylococcus epidermidis* (MRSE) and Methicillin Resistant *Staphylococcus saprophyticus* (MRSS) obtained from clinical pus samples of patients (Orthopaedic, postnatal, paediatric, ICU, female and male surgical wards) as well as other resistant Staphylococcus species isolated from blood and High vaginal swab from the MZCH Microbiology Laboratory sample repository. Out of the 23 tested isolates, 87% (20/23) of the clinical *Staphylococcus species* got lysed, and the majority of positive lysis outcomes were observed in isolates from pus samples, followed by blood and HVS isolates (table 2).

**Table 2.** Staphylophage host range analysis with a summary of sample type, participant’s clinical history, Staphylococcus species, and plaque formation outcome. Bacterial isolates obtained from pus, blood, and high vaginal swab (HVS) specimens were identified as methicillin-resistant Staphylococcus aureus (MRSA), methicillin-sensitive S. aureus (MSSA), methicillin-sensitive S. epidermidis (MSSE), methicillin-sensitive Staphylococcus spp. (MSSS), or methicillin-resistant Staphylococcus spp. (MRSS). Phage susceptibility was assessed using a plaque assay, where the presence of plaques indicated bacterial lysis by the bacteriophage. Phage titers are expressed as plaque-forming units per milliliter (PFU/mL). A dash (–) indicates that no plaques were observed and no phage titer was detected. PCS = post-caesarean section; CAVD = chronic abnormal vaginal discharge; ABD = abnormal vaginal discharge; HVS = high vaginal swab.

| Number | Sample type | Patient clinical information | Staph.Spp exposed to MRSA phage | Plaque formation | Titer (PFU/ml) |
| --- | --- | --- | --- | --- | --- |
| 1 | Pus | R. Leg amputation | MRSA | Lysis (plaques) | $100 \times 10^6$ |
| 2 | Pus | Post cesarean operation (PCS) | MRSA | Lysis (plaques) | $50 \times 10^3$ |
| 3 | Pus | Bone injury | MRSA | Lysis (plaques) | $21 \times 10^9$ |
| 4 | Pus | Amputated L.leg with implants | MSSE | Lysis (plaques) | $20 \times 10^6$ |
| 5 | Pus | Bowel obstruction | MRSA | No lysis (no plaques) | - |
| 6 | Pus | Hernia | MRSA | Lysis (plaques) | $18 \times 10^3$ |
| 7 | Pus | cholecystitis | MRSA | Lysis (plaques) | $31 \times 10^9$ |
| 8 | Pus | fractures | MRSA | Lysis (plaques) | $14 \times 10^{11}$ |
| 9 | Pus | Tendinitis | MSSA | Lysis (plaques) | $7 \times 10^9$ |
| 10 | Pus | arthritis | MRSA | Lysis (plaques) | $23 \times 10^{10}$ |
| 11 | Pus | Bone injury | MSSA | Lysis(plaques) | $7 \times 10^9$ |
| 12 | Blood | sepsis | MSSA | Lysis (plaques) | $60 \times 10^9$ |
| 13 | Blood | sepsis | MRSA | No lysis (no plaques) | - |
| 14 | Blood | Sepsis after PCS procedure | MRSA | Lysis (plaques) | $23 \times 10^9$ |
| 15 | Blood | sepsis | MRSA | Lysis (plaques) | $22 \times 10^8$ |
| 16 | Blood | sepsis | MSSE | Lysis (plaques) | $10 \times 10^6$ |
| 17 | Blood | sepsis | MSSA | Lysis (plaques) | $20 \times 10^{12}$ |
| 18 | Blood | sepsis | MSSA | No lysis (no lysis) | - |
| 19 | HVS | CAVD | MSSA | Lysis (plaques) | $23 \times 10^9$ |
| 20 | HVS | CAVD | MSSS | Lysis (plaques) | $19 \times 10^8$ |
| 21 | HVS | ABD | MRSS | Lysis (plaques) | $17 \times 10^6$ |
| 22 | HVS | PCS | MRSA | Lysis (plaques) | $12 \times 10^{12}$ |
| 23 | HVS | Chronic vaginal Discharge | MRSA | Lysis (plaques) | $7 \times 10^{10}$ |

### PH Stability Plot

To evaluate the stability and robustness of the isolated phage, its lytic activity was assessed under different physical parameters, including varying pH levels duration. The study observed Staphylophage was stable within a moderate PH range (around 6 to 11) with optimal activity typically seen between PH 6 and 10. The outcomes of measuring the phage infectivity after exposure to different PH, are summarized in **figure 5**.

**Figure 5.**
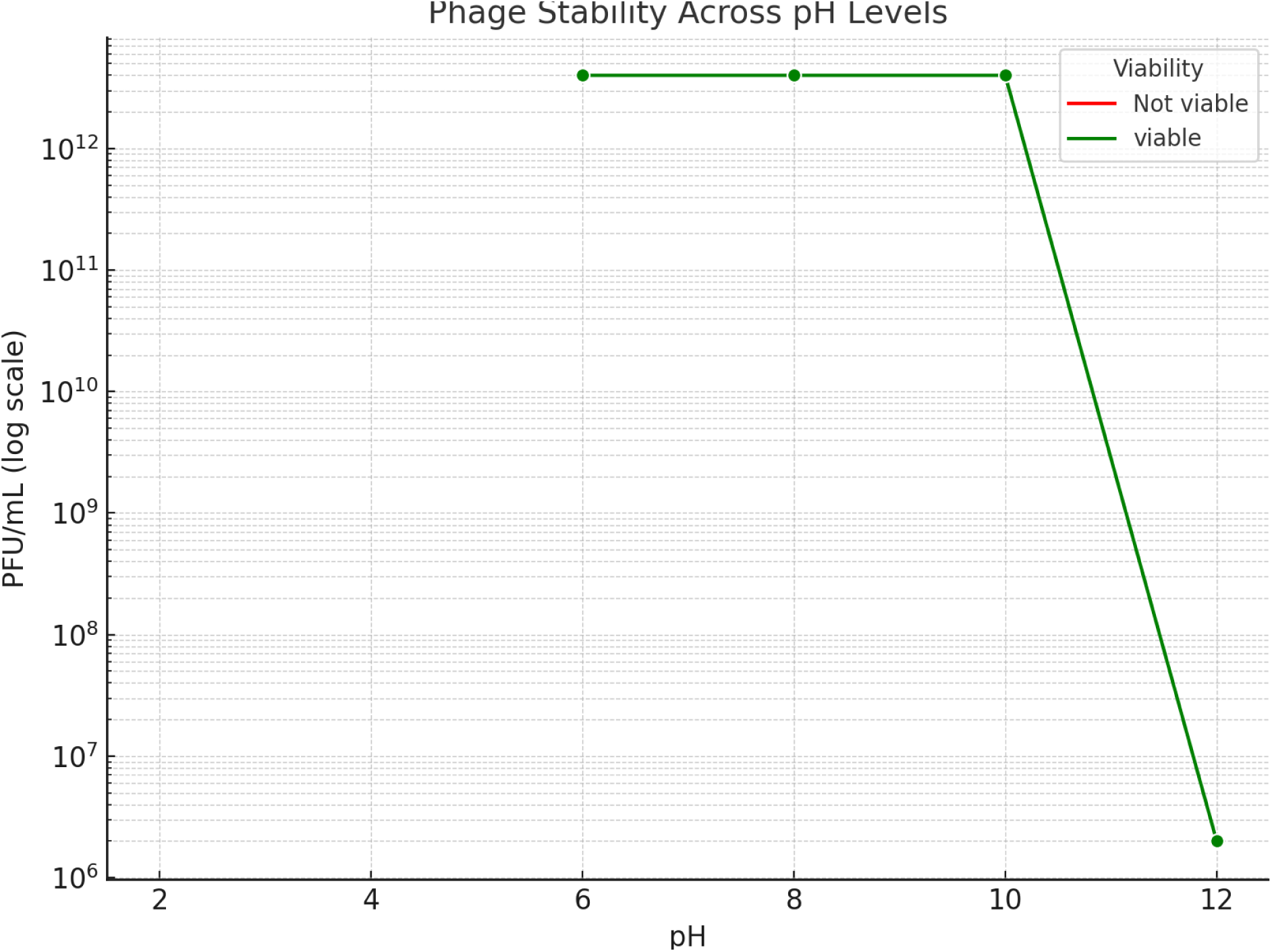
Stability of Staphylophage across different pH conditions. Maintaining phage viability is critical because it ensures that target Staphylophages remain active and infectious. Phage viability was evaluated by measuring plaque-forming units (PFU/mL) following exposure to pH values ranging from 2 to 12. Phage titers remained stable at approximately 3 × 10¹² PFU/mL under mildly acidic to neutral and moderately alkaline conditions (pH 6–10), indicating high stability within this pH range. In contrast, exposure to pH 12 resulted in a marked reduction in phage titer to approximately 2 × 10⁶ PFU/mL, demonstrating substantial loss of viability under highly alkaline conditions. Green markers indicate viable phage populations.

### Phage exposure to Ultraviolet light

Ultraviolet (UV) and Staphylophage testing involved using UV radiation to study and manipulate the isolated phage. The study measured effects of UV exposure to the isolated Staphylophage by observing phage lysis on the TSA plate. This was done by exposing phage to different doses of UV (5-60 minutes) and then measuring the surviving phage population. The figure 4.7 illustrates plaque formation (represents phages survival) after 30 minutes UV exposure and the summary of UV exposure for the rest of time. For easy interpretation and visualization, the results of UV exposure are further summarized in the **figure 6**.

**Figure 6.**
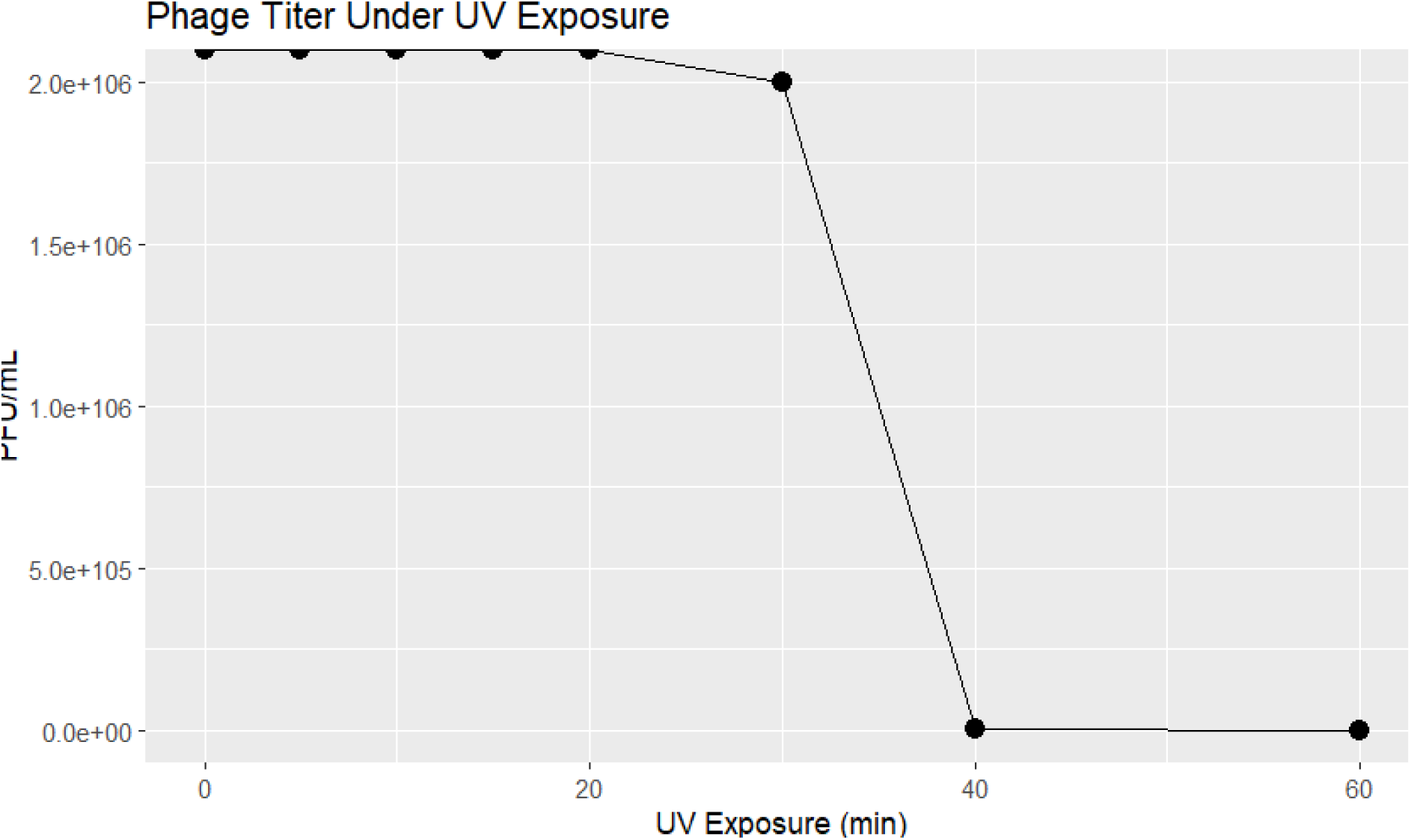
*Phage titer (PFU/mL) under UV exposure*. Phage titers remained stable for up to 30 minutes of UV exposure at wavelength of 254 nm, with concentrations maintained above 2.0 × 10⁶ PFU/ml. No significant reduction in phage viability was observed during this period, indicating resistance to short-term UV irradiation. However, a sharp decline in phage titer was recorded after 40 minutes of continuous exposure, with PFU/mL dropping to undetectable levels. No viable phage particles were recovered after 40, 50, and 60 minutes of exposure.

These results indicate a critical threshold for UV tolerance at or around the 30-minute mark. ANOVA analysis showed a statistically significant reduction in phage titer between the 30- and 40-minute time points (p < 0.01), confirming the susceptibility of phages to prolonged ultraviolet exposure.

## DISCUSSIONS

This prospective study of post-surgical wound infections conducted over a three-month period at Mzuzu Central Hospital revealed a high prevalence of methicillin-resistant *Staphylococcus aureus* (MRSA) among clinical isolates. Of the 56 bacterial isolates recovered from fifty wound swabs (50 participants), *S. aureus* accounted for 32.1% (18/56), of which 61.1% (11/18) were confirmed as methicillin-resistant. These findings highlight the significant burden of MRSA in post-surgical wound infections and underscore the need for effective infection prevention and control measures within the hospital setting (20). A lytic Staphylophage was successfully isolated from hospital sewage at Mzuzu Central Hospital and demonstrated potent lytic activity against MRSA isolates recovered from post-surgical wound infections. The successful isolation of an anti-MRSA Staphylophage from the local hospital environment suggests that bacteriophages may represent a promising alternative or complementary therapeutic approach for managing antibiotic-resistant wound infections (21). The high proportion of MRSA observed in this study is consistent with the growing global concern regarding antimicrobial resistance in healthcare settings (4). However, routine surveillance data generated through WHONET at Mzuzu Central Hospital indicate MRSA prevalence of approximately 20% during the 2023–2024 period. The higher prevalence observed in the present study may reflect differences in the study population, specimen type, sampling period, or the specific focus on post-surgical wound infections, which are often associated with increased exposure to healthcare-associated pathogens and antibiotic selection pressure.

The isolated Staphylophage from MZCH sewage exhibited 87% lytic activity against *Staphylococcus* species recovered from surgical wounds, blood, and high vaginal swab samples, indicating substantial bactericidal efficacy and a promising foundation for alternative antimicrobial therapy. These findings are consistent with similar phage isolation studies conducted in neighbouring African regions such as Ghana and Kenya, which have demonstrated the therapeutic potential of sewage-derived *MRSA phage* against MRSA strains (22).

Phage sensitivity assays revealed selective lytic activity in 87% of resistant clinical isolates, particularly in pus-derived MRSA samples, suggesting possible strain- or site-specific susceptibility patterns.

The isolated phage maintained thermal stability across a broad temperature range and showed robust viability at 4°C, implying that ultra-low storage temperatures (−80°C) may not be necessary for short-term applications. The isolated Staphylophage retained infectivity at pH 2– 12, temperatures from 4°C to -80°C, and up to 60 minutes of UV exposure, confirming their environmental robustness, which aligns with previous observations in African sewage-based phage studies (13).

The observed lytic activity likely reflects natural co-evolutionary adaptation between bacteriophages and hospital-associated bacterial populations within sewage ecosystems. The dual traits of broad host range and environmental stability suggest that such phages are optimized for persistence in dynamic and contamination-rich environments. The decline in phage viability beyond 30 minutes of UV exposure supports known susceptibilities of phage capsid proteins to ultraviolet degradation. Similar adaptive resilience and infection profiles have been documented in genomic and proteomic analyses of MRSA-targeting phages isolated from sewage and burn wound samples in Pakistan (23).

The results corroborate earlier reports from Egypt (Samir et al., 2022) and Iran (Elahi et al., 2021) that demonstrated successful recovery of MRSA-targeting phages from hospital effluents, reinforcing the concept of wastewater as a renewable source of therapeutic phages(13,17).

Likewise, they complement Rodwell et al. (2021) in Malawi, who reported efficient isolation of *Salmonella*-targeting phages from local sewage, highlighting the regional feasibility of phage bioprospecting (24). However, the lack of molecular characterization or genomic sequencing in the present study constrains direct phylogenetic comparison with other known staphylophage and underscores the need for detailed molecular analyses.

This research addressed a pressing local gap in antimicrobial resistance (AMR) management, demonstrating that both MRSA and MSSA strains yielded high phage titers (∼10¹³ PFU/mL). These findings imply enhanced phage susceptibility among more clinically virulent strains.

Conversely, *S. epidermidis* and *S. saprophyticus* exhibited lower PFU counts (10⁶–10⁷ PFU/mL), potentially reflecting weaker phage adsorption or replication efficiency. The biological variability among strains suggests underlying genotypic and phenotypic diversity influencing phage responsiveness.

The lack of technical and biological replicates due to resource limitations prevented computation of standard deviations or incorporation of error bars, restricting statistical interpretation.

Nonetheless, these exploratory data provide valuable preliminary evidence of phage viability trends across diverse environmental stresses. Beyond microbiology, these findings have public health and policy implications. The demonstrated feasibility of sourcing therapeutic phage locally supports efforts to reduce reliance on imported antibiotics and sheds light on hospital sewage as a potential reservoir for bioactive agents.

### Generalizations and Exceptions

A generalizable finding was that phages isolated from hospital sewage exhibited activity against both MRSA and Methicillin-Sensitive *S. aureus* (MSSA), as well as resistant Coagulase-Negative *Staphylococcus species*. However, exceptions were noted in the variability of lytic activity across sample types, and one isolate demonstrated pan-drug resistance (PDR), for which phage lysis was not tested, leaving open questions about phage efficacy in extreme resistance scenarios.

In this study, phage activity (PFU/mL) was measured across a variety of experimental conditions including storage temperatures, pH levels, and UV exposure durations. Due to logistical and resource constraints during laboratory experimentation, most conditions were assessed using single measurements without technical or biological replicates. Consequently, it was not possible to compute standard deviations or include error bars in the graphical representations. While this limits the ability to quantify variability or assess statistical confidence across replicates, the results still provide useful exploratory insights into the trends and comparative effects of these stress conditions on phage viability.

### Multiple Hypotheses and Interpretive Caution

While the data supports phage efficacy, alternative explanations must be considered. The observed lytic activity might not generalize to all MRSA strains, especially those from different geographical or clinical contexts. Moreover, phage-host specificity could evolve over time, potentially reducing long-term efficacy. The role of co-infecting organisms and biofilm. Formation in wound sites may also influence phage effectiveness.

The biological relevance of these results lies in their implications for phage therapy potential. For instance, the observed variability in titers suggests that certain strains, particularly MRSA, may exhibit differential susceptibility to phage infection— highlighting potential for targeted phage therapy. The positive Spearman correlation between plaque formation and PFU/mL further supports the functional integrity and lytic potential of the phages used, indicating that measurable titers correspond to observable biological effects on bacterial lawns. Even in the absence of statistical significance due to limited replicates, these trends provide preliminary evidence of strain-specific responses and the feasibility of phage survival under harsh conditions such as UV and acidic pH exposure. This lays the groundwork for more comprehensive, replicated studies that could validate the therapeutic promise of these lytic phages in clinical settings, especially for antibiotic-resistant infections.

Ultimately, the integration of statistical outcomes with biological context enhances the interpretability and applied value of the data, bridging bench-level observations with real-world therapeutic implications. Thus, while phage therapy shows promise, it should be approached as a complementary—not replacement—strategy alongside antibiotics. Ongoing monitoring, host range studies, and resistance evolution tracking are essential to validate and refine this therapeutic approach.

### New Knowledge and Significance

This study provides the first documented evidence of MRSA-specific phage isolation in Malawi. It establishes baseline knowledge of MRSA prevalence in surgical wards at MZCH and confirms hospital sewage as a viable source of therapeutic phages. It also contributes to understanding phage stability across environmental stressors. Collectively, these findings pave the way for integrative AMR mitigation strategies rooted in locally relevant data.

The implications for patient care, environmental microbiology, and antimicrobial stewardship underscore why these findings matter: they address an urgent health crisis with an innovative, sustainable, and context-sensitive approach.

## CONCLUSIONS AND RECOMMENDATIONS

### Conclusions

This study presents strong evidence that lytic bacteriophages isolated from hospital sewage at Mzuzu Central Hospital can effectively target MRSA strains derived from postsurgical wound infections. The most important outcome of this work is the demonstration that locally sourced phages are not only viable and stable but also exhibit significant lytic activity against clinically relevant resistant bacteria.

If there is one finding to be remembered, it is this: hospital sewage in Malawi is a viable and untapped source of therapeutic bacteriophages capable of combating antimicrobial-resistant pathogens like MRSA. The study managed to isolate a phage (Staphylophage) capable of infecting 87 % of clinically isolated resistant bacteria from postsurgical wards at MZCH in host range analysis.

The investigation addressed a critical gap by confirming that 61.1% of *S.aureus* (11/18) isolates from surgical patients were methicillin-resistant and that lytic phages isolated from environmental samples could target these strains. This study introduces new insights into the feasibility of phage therapy in Malawi and offers preliminary data supporting the stability and broad activity of phages under various environmental conditions.

These findings have broader implications for antimicrobial resistance management, pointing to an alternative treatment pathway that is context-appropriate, sustainable, and scalable. Phage therapy, if further validated, could enhance local health systems’ capacity to manage resistant infections without over-reliance on expensive or inaccessible antibiotics.

### Recommendations

The identification of elevated resistance rates in the current study emphasizes the necessity for greater antimicrobial stewardship to halt the development and escalation of AMR. Strengthening Antimicrobial Stewardship Programs by implementing more stringent antimicrobial stewardship programs. These programs must emphasize the proper use of antibiotics, according to existing resistance patterns, and reducing unnecessary prescriptions to avoid further resistance. Also, surveillance of the trends in AMR should be done regularly to ensure that healthcare providers are properly informed about the resistance patterns in pathogens. It is recommended that health policymakers integrate phage research and development into Malawi’s national antimicrobial resistance (AMR) action strategy. Further study on MRSA sequence and phage to determine regions of interest for possibility of producing antibiotics to treat MRSA. Given the demonstrated efficacy and environmental robustness of phages isolated from hospital sewage, these agents should be considered in national-level interventions, especially in high-risk surgical settings. Furthermore, genomic and molecular characterization of these phages is essential to determine their safety, taxonomy, and therapeutic viability. Sequencing will also help identify possible virulence or resistance genes that might be present. Future studies should explore the use of these phages in animal models, followed by human clinical trials, to establish safety and efficacy under in vivo conditions. The scope of phage surveillance should also be expanded to include additional referral hospitals in Malawi to understand regional variability in MRSA strains and corresponding phage efficacy. Because single phages may not always provide broad protection, development of multi-phage cocktails should be pursued to enhance treatment outcomes and reduce the emergence of phage resistance. Additionally, collaborations between academic institutions, biotechnology firms, and hospitals are needed to build capacity for phage research, formulation, and local production. Environmental monitoring of hospital sewage could also serve as a dual-purpose tool for detecting resistant pathogens and continuously sourcing novel phages. Overall, these recommendations support the establishment of a sustainable, local phage therapy framework that complements existing antibiotic strategies.

## Data Availability

All data produced in the present study are available upon reasonable request to the authors.

## LIST OF ABBREVATIONS

MOH: Ministry of Health
AMR: Antimicrobial Resistance
MUSTREC: Malawi University of Science and Technology Research Ethical Committee
MRSA: Methicillin Resistant *Staphylococcus aureus*
UV: Ultra Violet light
MZUNI: Mzuzu University
PHIM: Public Health Institute of Malawi
MZCH: Mzuzu Central Hospital
MSSE: Methicillin Susceptible *Staphylococcus epidermidis*
MRSS: Methicillin Resistant *Staphylococcus saprophyticus*
EUCAST: European Union Committee for Antimicrobial Susceptibility Testing
MUST: Malawi University of Science and Technology
CLSI: Clinical and Laboratory Standards Institute
AMS: Antimicrobial Stewardship

## DECLARATION

### ETHICS APPROVAL AND CONSENT TO PARTICIPATE

#### Ethical Considerations

Prior to the initiation of the study, ethical approval was sought from Malawi University of Science and Technology Research Ethics Committee (MUSTREC), reference number: P.07/2024/159. MUSTREC is registered with the USA Office for Human Research Protections (OHRP) as an international IRB (IRB Number IRB00012588 FWA00029630). The permission to conduct the study at Mzuzu Central Hospital (MZCH) was sought from MZCH Committee) (MZCHRC) through the hospital director. Both verbal and formal written consent was obtained in accordance with institutional guidelines. Study aims and procedures were initially explained to each participant. Each participant had the right to decline or participate without coercion or any undue incentives. No identifiable patient information was obtained from the study participants.

### CONSENT FOR PUBLICATION

Not applicable

### AVAILABILITY OF DATA AND MATERIAL

Raw data and material are readily available if required at any time.

